# People living alone with neurodegenerative conditions: a scoping review protocol

**DOI:** 10.1101/2025.03.12.25323814

**Authors:** Anthony Martyr, Maria Caulfield, Catherine Charlwood, Laura D. Gamble, Matthew Prina, Jan R. Oyebode, Claire Hulme, Linda Clare

## Abstract

**Introduction:** People living alone with neurodegenerative conditions face unique difficulties in maintaining independence and accessing appropriate health and social care support. Consolidating current understanding regarding these unique difficulties would better inform health and social care services and enable more tailored and appropriate service delivery. The proposed scoping review will summarise evidence from studies that provide evidence about people with dementia, Parkinson’s disease, Huntington’s disease, or motor neurone disease who live alone.

**Methods and Analysis:** This scoping review protocol uses well-established methodology outlined by the Preferred Reporting Items for Systematic review and Meta-Analysis and the Joanna Briggs Institute. Relevant publications will be searched using PubMed, Web of Science Core Collection, CINAHL and AgeLine via EBSCOhost, and EMBASE, PsycInfo, and Social Policy and Practice via Ovid. Grey literature will be searched via Google looking specifically for pdf documents. No date restrictions will be applied to the searches. For research publications a two-stage screening approach will be undertaken. The first stage will involve screening titles and abstracts for relevant literature on people with neurodegenerative conditions living alone in the community. The second stage will involve full text screening of selected articles. For grey literature the first 20 pdfs per website identified in Google will be downloaded and screened. Summary data will be extracted from publications selected for inclusion. Data synthesis will involve tabulating study characteristics and findings and preparing narrative summaries to identify commonalities, gaps, and areas for future research.

**Ethics and Dissemination:** Ethical approval is not required for this review. Consultation with people with lived experience, stakeholders and experts linked with the National Institute for Health and Care Research (NIHR) Policy Research Unit in Dementia and Neurodegeneration University of Exeter (DeNPRU Exeter), will help to ensure the relevance and applicability of findings. Dissemination will include a policy report and a peer-reviewed publication aimed at informing policy, practice, and improving support services for people living alone with neurodegenerative conditions.

**Article Summary:** Strengths and limitations of this study

- The use of a rigorous and well-established methodological framework will enhance the quality and structure of the review.
- A comprehensive search will be conducted in 7 databases will maximise identification of all possible records that meet the inclusion criteria of the review.
- The inclusion of grey literature will complement the research literature by providing detail about guidance being given to people living with and people caring for neurodegenerative conditions.
- A limitation is that the literature search will be restricted to articles published in English. This increases the risk that relevant information will be missed.

## Background

Since the 1960s the proportion of adults living alone has steadily increased (Snell, 2017). In the United Kingdom (UK), census data show that in 2021 30.1% of adults lived alone (Office for National Statistics, 2023b), and this proportion is largely consistent with other wealthy nations in Europe and North America (Snell, 2017). Living alone has become more common due to societal changes and shifts in lifestyle preferences. Factors such as increased urbanisation, delayed marriage, higher divorce rates, and a focus on individualism and personal autonomy have all contributed to the rise in the number of adults living alone (Esteve et al., 2020). These factors largely affect younger or middle-aged adults whereas different factors are likely to be more salient for older adults. This is important as living alone is increasing most rapidly among older adults (Eurostat, 2020; Office for National Statistics, 2023a), and it is especially common in older women (Abell et al., 2021), who are more frequently widowed than men (Office for National Statistics, 2024).

As the proportion of people living alone continues to increase it is to be expected that there will be a concomitant increase in the number of people living alone in the community with a neurodegenerative condition. The four most common neurodegenerative conditions are Alzheimer’s disease and other types of dementia, Parkinson’s disease, Huntington’s disease, and motor neurone disease (Fratiglioni et al., 2009), including amyotrophic lateral sclerosis or Lou Gehrig’s disease. Rarer neurodegenerative conditions include prion diseases like Creutzfeldt-Jakob disease, multiple system atrophy, progressive supranuclear palsy, and ataxia. Most of these conditions are associated with increased age and are more prevalent in people over 65, though Huntington’s disease, motor neurone disease, and some types of dementia often appear in people aged under 65. It is estimated that worldwide there are over 55 million people living with dementia (Long et al., 2023), more than 10.9 million people living with Parkinson’s disease (Steinmetz et al., 2024), around 400,000 people living with Huntington’s disease (Medina et al., 2022), and around 260,000 people living with motor neurone disease (Steinmetz et al., 2024). Precise population-based estimates of the number of people living alone in the community with these conditions in the UK are lacking. Gould et al. (2015) estimated that, in the United States of America (USA) in 2011, 13% of people with dementia were living alone in the community. Where research studies have recruited samples that include people with dementia living alone or with others in the community, the proportion of people living alone varies considerably, from 18.5% in a cohort study recruited in Britain (Clare et al., 2024; Clare et al., 2020) to 51% in a German sample (Eichler et al., 2016). These differences are likely to be due to variation in sampling methods. One online study that included nearly 5,500 people with Parkinson’s disease and recruited globally, though around 80% were from the USA, reported that 12% lived alone (Brown et al., 2020).

People living alone with neurodegenerative conditions may require more support to maintain a safe and fulfilling lifestyle than those living with others. Most people living alone with neurodegenerative conditions have access to varying degrees of help and support from unpaid or informal carers, typically family or friends (Clare et al., 2020; Gould et al., 2015; Nwabuobi et al., 2019). Indeed, it is widely assumed that a family carer is available to provide support to the person if needed and act as a source of information for health and social care professionals. Where a family carer is not available to assist, some studies suggest people with dementia may have difficulties in engaging and interacting with services and may find services unresponsive (Portacolone et al., 2019). Additionally, people with Parkinson’s disease find services difficult to access (Tod et al., 2016). These challenges may increase as the severity of the condition progresses. Over time, the service use of people living with these conditions tends to rise (Parkinson’s UK, 2016), with costs for people with dementia living alone being around 35% higher over two years compared to those living with others (Henderson et al., 2022). As the number of people living alone with neurodegenerative conditions increases this will inevitably escalate the demands placed on health care service providers and stretch service capacity.

Evidence suggests that people who live alone with neurodegenerative conditions are at an increased risk of numerous adverse outcomes. These include increased social isolation, increased loneliness, lower satisfaction with life, greater exploitation, higher incidence of accidental injury, elevated risk of malnutrition, increased psychological distress, greater self-neglect, and a higher rate of admission to residential care (Cao et al., 2021; Clare et al., 2024; Gould et al., 2015; Tod et al., 2016). Therefore, as the number of people living alone with neurodegenerative conditions continues to rise, it is important to identify the unique challenges to living alone with these conditions. Furthermore, it is essential to develop comprehensive strategies within healthcare systems to meet their needs, as addressing these needs not only enhances quality of life but also reduces the burden on healthcare systems through for example preventing crises and avoidable hospital admissions. This, in turn, promotes more efficient resource allocation and improved outcomes.

## Study Rationale

It is important to consider how people with neurodegenerative conditions cope when living alone, especially as the number of people living alone with a neurodegenerative condition is increasing and family carers are not always available. Understanding the diverse needs and challenges associated with living alone with each condition can inform policy and, in turn, influence how services are structured, planned, and delivered. This applies to both the voluntary sector and health and social care provision for people living with a neurodegenerative condition, as well as practitioner training and skills development.

While most research on this topic focuses on dementia, by considering this issue across a range of neurodegenerative conditions we aim to highlight differences and similarities in care delivery and needs. This can help to identify good practice and generate recommendations for improvement in care and support services for people living alone with these conditions. To do this we need to understand the needs and challenges that people experience when living alone, gain an overview of how services are provided, identify gaps in care coordination and integration, and provide recommendations for training and education in the health and social care workforce. Synthesising available evidence will result in recommendations that can inform the development of policies and services tailored to meet the diverse needs of people with neurodegenerative conditions who live alone.

To date, there have been no comprehensive systematic reviews or scoping reviews analysing the extant literature on people who live alone with neurodegenerative conditions. Using a scoping method allows for a thorough and systematic review process while also providing flexibility to incorporate various types of studies, such as quantitative, qualitative, mixed methods, and case studies. Conducting a scoping review on this topic will provide insights into the challenges, coping mechanisms, and support systems of people living alone with neurodegenerative conditions. The review will help to identify gaps in our understanding and where future research should be directed. It will enable us to inform policy and practice and identify key areas of health and social care where changes would provide the most benefit.

### Study Objectives

The aim of this scoping review is to systematically assess both research and grey literature concerning people with neurodegenerative conditions who live alone in the community. In the review we will:

- Identify the characteristics of people living alone, their experiences, needs, challenges, resource use and associated costs to the health and social care sectors, individuals, and families, and how these change over time.
- Evaluate current practice from practice guidelines, information for practitioners, and practical advice for people with different neurodegenerative conditions. This will show what information is available, what practitioners are doing, and what might constitute good or best practice.
- Consider inequities experienced by people with neurodegenerative conditions who live alone relative to those living with others, which may include issues related to social isolation and exclusion, access to health and social care services, and support from carers.

We will use the evidence synthesised in the review to:

- Generate recommendations for policy and practice.
- Highlight any gaps in knowledge that may represent areas for future investigation.

## Methods and analysis

The approach to be taken is based on the protocol methodology outlined in Preferred Reporting Items for Systematic review and Meta-Analysis Protocols (PRISMA-P) guidance (Moher et al., 2015; Shamseer et al., 2015), and Joanna Briggs Institute best practice guidance for scoping reviews (Peters et al., 2022).

Eligibility criteria

The following types of sources will be included:

- Studies that report information about people with neurodegenerative conditions who live alone; this includes both studies that focus solely on people living alone and studies that compare people living alone with people living with others.
- Studies that focus on people with dementia, Parkinson’s disease, Huntington’s disease, or motor neurone disease. Studies reporting on rarer neurodegenerative conditions will also be included. It is anticipated that most studies will involve people with dementia, with fewer studies covering other neurodegenerative conditions.
- Studies where samples are described as having ‘cognitive impairment’ as it is likely that they will include people who have dementia but lack a formal diagnosis. These will typically involve older people with undiagnosed dementia or significant cognitive impairment who rarely contact health services. Such individuals can only be reported descriptively as having ‘cognitive impairment’ and those without a definite diagnosis may be in the greatest need of support, making policy direction most urgently required.
- Studies that report on mixed diagnostic groups where cognitive impairment is a factor (e.g., studies that include those with neurodegenerative conditions but also people with stroke, head injury, or other acute insults). These studies will be included if one of the following conditions is met:

1. they provide information about people with relevant conditions separately.
2. if conditions are not reported separately, at least 80% of the sample must include people with relevant neurodegenerative conditions, provided other inclusion criteria are met.
- Grey literature that offers support or advice to people living alone with neurodegenerative conditions, their carers, or health and social care professionals.
- Grey literature that addresses topics of relevance to people living alone with neurodegenerative conditions, such as managing autonomy and risk, loneliness, ensuring home safety, planning for future care, etc.

The following types of sources will be excluded:

- Studies focusing on people diagnosed with mild cognitive impairment. These people do not have dementia and most people diagnosed with mild cognitive impairment either remain stable or revert to normal cognitive functioning for their age (Ganguli et al., 2019). This group differs from the above ‘cognitive impairment’ group as mild cognitive impairment is a specific diagnosis.
- Studies focusing on people diagnosed with multiple sclerosis as this is considered an autoimmune disease rather than a neurodegenerative condition.
- Studies focusing on people classified as having “cognitive impairment not dementia” or “CIND” as, while these people have cognitive impairment, dementia has specifically been excluded.
- Studies that report on people with cognitive impairment due to head injury, stroke, or other acute insults.
- Grey literature that refers to living alone within the document but lacks substantial commentary, advice, or relevance to the experience or needs of people living alone with a neurodegenerative condition and/or their carers.

Other exclusion criteria are:

- Articles written in languages other than English.
- Articles for which full text versions are unavailable.
- Reviews, editorials, letters, and opinion pieces.
- Published conference abstracts; these will be used to find subsequently published articles where available. When articles are found via this route, they will be included in the ‘identified via other sources’ section of the PRISMA flowchart.
- Studies describing mixed samples that do not meet the inclusion criteria will be excluded. This includes studies where:

- The sample includes people with one or more of the included conditions (e.g., dementia) along with one or more conditions from the excluded groups (e.g., mild cognitive impairment), but less than 20% of the sample comprises people with relevant conditions, and data are not reported separately for different conditions.

## Information sources

Relevant studies will be identified by searching seven electronic databases covering published research literature: PubMed, Web of Science Core Collection, CINAHL and AgeLine via EBSCOhost, and EMBASE, PsycInfo, and Social Policy and Practice via Ovid. There will not be any date restriction on these searches.

Grey literature including reports, information booklets, advice leaflets, practice guidelines, etc., will be searched using advanced Google search parameters that will be applied to 76 preselected websites; a list of these 76 websites can be found in supplementary material. The 76 websites have been selected as they are either the main English language condition-specific websites for dementia, Parkinson’s disease, Huntington’s disease, or motor neurone disease, hosted by organisations in the UK, the USA, Canada, Europe, or Australia, or websites of widely known global health organisations or health and social care think tanks. Grey literature searches will be restricted to pdf documents only. As the most applicable pdfs will be returned at the top of the search engine results page, the first 20 pdfs that Google returns for each website will be downloaded and screened. Where the search returns fewer than 20 pdfs, all pdfs that are returned will be screened.

## Search strategy

The search terms to be used were collated by screening recent reviews of other conditions that have used synonyms for living alone. Search terms will be restricted to English language publications when databases permit using this restriction. No other restrictions will be applied.

Search terms comprise two components: a string comprising terms related to living alone and a neurodegenerative condition string.

Living alone string terms are:

- Living alone OR Live* alone OR Single-living OR One-person household OR Singlehood OR Single people OR Single person OR Single men OR Single women OR solo

Neurodegenerative condition string terms are:

- dement* OR Alzheimer* OR Parkinson* OR Lewy OR Fronto* OR Huntington* OR Chorea OR amyotrophic lateral sclerosis OR ALS OR motor neuron* disease OR MND OR progressive muscular atrophy OR Gehrig OR neurodegen* OR neurolog* OR cognitiv* impair*

Here is an example search string for Ovid:

((dement* or Alzheimer* or Parkinson* or Lewy or Fronto* or Huntington* or Chorea or amyotrophic lateral sclerosis or ALS or motor neuron* disease or MND or progressive muscular atrophy or Gehrig or neurodegen* or neurolog* or cognitiv* impair*) and (Living alone or Live* alone or Single-living or One-person household or Singlehood or Single people or Single person or Single men or Single women or solo)).ti,ab.

These search strings will be adapted when searching for grey literature. Different search terms will be used for condition-specific and non-condition-specific websites.

For condition-specific websites, the search will use these living alone synonyms only:

- “Living alone” OR “Live alone” OR “Lives alone” OR “Lived alone”

An example search string for a condition-specific website is:

- site:alzheimers.org.uk “Living alone” OR “Live alone” OR “Lives alone” OR “Lived alone” filetype:pdf

For non-condition-specific websites, searches will be conducted using each of these four search terms separately, combined with an adapted list of the condition terms used in the published literature search described above. An example search string is:

- site:who.int “living alone” dementia OR demented OR Alzheimer OR Parkinson OR Lewy OR Fronto OR Huntington OR Chorea OR “amyotrophic lateral sclerosis” OR ALS OR “motor neuron disease” OR MND OR “progressive muscular atrophy” OR Gehrig OR neurodegenerative OR neurodegeneration OR “cognitive impairment OR “cognitively impaired” filetype:pdf

## Data management

Literature search results will be imported into EndNote. Duplicates will be removed using a series of duplicate searches within EndNote. A final manual screen for any additional duplicates will be conducted. Title, abstract, and full text screening stages will be undertaken within EndNote. For grey literature searches, pdfs will be downloaded into folders, one folder for each website searched. Details of each search will be recorded in an Excel spreadsheet including the search date, the name of the researcher who conducted the search, the search string(s) used for each website, the number of pdfs returned in the search engine results page, the number of duplicates manually removed, and the number of pdfs extracted for screening.

## Selection process

The selection processes for research and grey literature are described separately below. Reasons for exclusion at the full text stage will be recorded and reported. Records of all searches will be recorded and summarised using PRISMA guidelines and recommendations for reporting.

## Research literature

The study selection process will comprise two levels of screening. Following removal of duplicates, titles and abstracts of all returned records will be screened by two reviewers working independently. Any records that are deemed potentially relevant by either or both reviewers will be taken forward to the next stage. In this second stage, the full text of each record will be downloaded and examined against eligibility criteria for inclusion in the review. Full texts will be screened for eligibility by one researcher. To check for consistency, 20% of these will be screened by a second researcher, blinded to the decision of the first researcher. Where it is not clear whether articles should be included, decisions about eligibility will be made via group discussion involving a third reviewer. Neither journal titles nor author names will be blinded during any screening stage.

Study selection will focus on articles published in English for which full text is available. Quantitative, qualitative, mixed method, and case studies will be considered. Where randomised controlled trials are included, only information from the baseline analysis will be used, except where the trial reports intervention effects separately for people living alone and people living with others, in which case data from follow-up assessments will also be used.

Multiple articles that use data from the same study will be identified and collated to avoid duplication of findings. Where the same variables are included in multiple articles from the same study, preference will be given to the article that reports the largest sample size. Where sample sizes are identical and where the same variables are included, data from the most recent article to be published from the same study will be used.

## Grey literature

For grey literature, a similar procedure will be used. As items of grey literature may not contain abstracts, the full text of all pdfs that are downloaded will be screened following the full text screening procedure outlined above for research articles. Any research articles or published conference abstracts identified in the grey literature search will be cross-checked with the research literature search and either excluded if already found or screened as described above for research literature. For recording purposes any articles identified in this way will be included in the ‘identified via other sources’ section of the PRISMA flowchart.

## Data collection process and data items

Data extracted from research and grey literature will be recorded using Excel spreadsheets.

## Research literature

Study characteristics that will be extracted are:

- Information about the study: first author name, publication year, study methodology, date that study data were collected, study name or funding grant number to allow identification of multiple articles that use data from the same study, and country or countries where data collection took place.
- Information about the sample or population: participant age, sex, education, ethnicity, diagnosis, diagnostic criteria used, severity – either cognitive screening mean score or other condition-specific severity metric such as Hoehn and Yahr stage and/or Unified Parkinson’s Disease Rating Scale score, and/or number of people in severity bands, and number and/or proportion of people with a neurodegenerative condition.
- Information about living situation: number and/or proportion of people living alone, and other included groups that are not living alone.
- Information about variables relevant to the research questions regarding living alone.
- Any recommendations for policy or practice proposed by the study authors.

The following procedures will be followed for quantitative and qualitative findings:

- For quantitative studies, data will be extracted for all variables relevant to living alone; this will comprise both data relating to findings and summary statistics. Where comparisons are made, relevant data will be extracted for people living alone and for people not living alone.
- For qualitative studies overarching themes and subthemes, representative quotations where applicable, and main findings related to living alone, will be extracted.

Studies will not be excluded where some of this information is missing; where this is the case, data that are missing will be clearly identified.

Grey literature

For grey literature we will extract:

- Information about the document: publication year, organisation on behalf of which the publication was produced, author, document type, country of origin, specific context surrounding discussions on living alone.
- Details of the processes used to gather information, such as surveys, interviews, and consultation, etc., where relevant.
- Information provided in the document about living alone with a neurodegenerative condition.
- Advice, support, guidance, or recommendations provided in the document for policymakers, practitioners, or people living with the condition.
- Any recommendations made to improve treatment and care for people living alone with neurodegenerative conditions.
- Any other potentially relevant information.

## Data synthesis

Data will be presented in tables that describe the study characteristics and the findings for each included study. There will be separate tables for quantitative and qualitative research literature and for grey literature. Mixed methods studies will be tabulated in applicable tables where relevant, or in a separate table. The proportion of people living alone with each condition will be calculated and presented. A narrative summary will accompany the table of results and will describe how the results relate to the review aim and research questions.

Findings will be grouped and described in the results section.

The Campbell and Cochrane Economic Methods Group Brief Economic Commentary framework (Shemilt et al., 2019) will be used to summarise the main findings of studies reporting health economic data.

Results will be reported in line with the PRISMA scoping review extension (Tricco et al., 2018).

## Stakeholder consultation

Stakeholders and people with lived experience have been involved in developing plans for this review. Once review findings are available, further consultation will be undertaken to examine these in relation to lived experience and practice and help to ensure resulting recommendations are relevant and realistic. Stakeholder consultation will draw on the expertise within the DeNPRU Exeter Knowledge Exchange Community, which includes experts by experience alongside other stakeholders. Additional specific expertise will be sought where needed.

## Dissemination and ethics

The review will be written up for publication. The findings will form part of a wider project on the topic conducted by the National Institute of Health and Care Research Policy Research Unit in Dementia and Neurodegeneration University of Exeter (DeNPRU Exeter). Ethical approval is not required for this study.

## Funding statement

This work was supported by National Institute for Health and Care Research (NIHR) Policy Research Unit in Dementia and Neurodegeneration (University of Exeter), reference NIHR206120. The grant was awarded to Linda Clare, Martin Knapp, Adelina Comas Herrera, David Burn, Louise Robinson, John-Paul Taylor, Matthew Prina, Jan Oyebode, Catherine Quinn, Sahdia Parveen, Claire Hulme, Kaarin Anstey, Rachael Litherland, Jeffrey Fox, & Julia Burton. This report is independent research supported by the National Institute for Health and Care Research Applied Research Collaboration South-West Peninsula. The views expressed in this publication are those of the authors and not necessarily those of the NIHR, the Department of Health and Social Care, or the National Health Service. The support of NIHR is gratefully acknowledged.

## Data Availability

No additional data are available.

## Acknowledgements

We are grateful to the DeNPRU Exeter Operations Manager Sara Hayes, members of the DeNPRU Exeter involvement group, and the Project Advisory Group for their support throughout the study. We would like to thank Rachael Litherland for organising the involvement group, and members Jane Ward and Nicholas Wrigley for their help and direction on the DeNPRU Exeter living alone project. For the purpose of open access, the authors have applied a Creative Commons Attribution (CC BY) licence to any Author Accepted Manuscript version arising.

## Author contributions

Drafting of article: Anthony Martyr. Research question ideation: Linda Clare. Funding acquisition: Linda Clare, Matthew Prina, Jan R. Oyebode, Claire Hulme. All authors contributed to the critical revision of the article and approved the version to be published.

## Competing interests

None declared.

Patient consent for publication

Not required.

Provenance and peer review

Not commissioned; externally peer reviewed.

## Data sharing statement

No additional data are available.

## Notes

### Competing Interest Statement

The authors have declared no competing interest.

