## Supplementary Material for "People living alone with neurodegenerative conditions: a scoping review protocol"

Alphabetical list of websites included in grey literature searches

a) Condition specific websites

| Number | Organisation | Website used in the search |
| --- | --- | --- |
| 1 | All-Party Parliamentary Group on Dementia (UK) | alzheimers.org.uk/about-us/policy-and-influencing/all-party-parliamentary-group-dementia |
| 2 | All-Party Parliamentary Group on MND (UK) | mndassociation.org/get-involved/campaigning/all-party-parliamentary-group-appg-on-mnd |
| 3 | All-Party Parliamentary Group on Parkinson's (UK) | parkinsons.org.uk/get-involved/parkinsons-uk-parliament |
| 4 | All-Party Parliamentary Group on Rare, Genetic and Undiagnosed Conditions (UK) | geneticalliance.org.uk/appg/ |
| 5 | ALS Society of Canada | als.ca |
| 6 | ALS Therapy Development Institute (USA) | als.net |
| 7 | Alzheimer Europe | alzheimer-europe.org |
| 8 | Alzheimer Scotland | alzscot.org |
| 9 | Alzheimer Society of Canada | alzheimer.ca |
| 10 | Alzheimer's Association (USA) | alz.org |
| 11 | Alzheimer's Disease International | alzint.org |
| 12 | Alzheimer's Research UK | alzheimersresearchuk.org |
| 13 | Alzheimer's Society (UK) | alzheimers.org.uk |
| 14 | Alzheimer's Society of Ireland | alzheimer.ie |
| 15 | American Parkinson Disease Association | apdaparkinson.org |
| 16 | Davis Phinney Foundation for Parkinson's (USA) | davisphinneyfoundation.org |
| 17 | Dementia Australia | dementia.org.au |
| 18 | Dementia Society of America | dementiasociety.org |
| 19 | Dementia UK | dementiauk.org |
| 20 | European Huntington Association | eurohuntington.org |
| 21 | European Huntington's Disease Network | ehdn.org |
| 22 | Fight MND (Australia) | fightmnd.org.au |
| 23 | Huntington Society of Canada | huntingtonsociety.ca |
| 24 | Huntington's Disease Association (UK) | hda.org.uk |
| 25 | Huntington's Disease Society of America | hdsa.org |
| 26 | Huntington's Disease Tasmania | huntingtonstasmania.org.au |
| 27 | Huntington's Disease Youth Organisation (USA & UK) | hdyo.org |
| 28 | Huntington's Victoria (Australia) | huntingtonsvic.org.au |
| 29 | Huntington's Western Australia | huntingtonswa.org.au |
| 30 | International Alliance of ALS/MND Associations | als-mnd.org |
| 31 | International Huntington's Disease Association | huntington-disease.org |
| 32 | International Parkinson and Movement Disorder Society | movementdisorders.org |
| 33 | Irish Motor Neurone Disease Association | imnda.ie |
| 34 | Les Turner ALS Foundation (USA) | lesturnerals.org |
| 35 | Lewy Body Dementia Association (USA) | lbda.org |
| 36 | Lewy Body Dementia Canada | lewybodydementia.ca |
| 37 | Lewy Body Ireland | lewybodyireland.org |
| 38 | Lewy Body Society (UK) | lewybody.org |
| 39 | MND Scotland | mndscotland.org.uk |
| 40 | Motor Neurone Disease Association (UK) | mndassociation.org |
| 41 | National Institute of Neurological Disorders and Stroke (USA) | ninds.nih.gov |
| 42 | New South Wales HD Association (Australia) | huntingtonsnsw.org.au |
| 43 | Parkinson Canada | parkinson.ca |
| 44 | Parkinson's Australia | parkinsons.org.au |
| 45 | Parkinson's Europe | parkinsonseurope.org |
| 46 | Parkinson's Foundation (USA) | parkinson.org |
| 47 | Parkinson's Ireland | parkinsons.ie |
| 48 | Parkinson's UK | parkinsons.org.uk |
| 49 | Queensland HD Association (Australia) | huntingtonsqld.org.au |
| 50 | Rare Dementia Support (UK) | raredementiasupport.org |
| 51 | Scottish Huntington's Association | hdscotland.org |
| 52 | Shake It Up Foundation (Australia) | shakeitup.org.au |
| 53 | South Australia and Northern Territory HD Association | huntingtonssant.org.au |
| 54 | The ALS Association (USA) | als.org |
| 55 | World Dementia Council | worlddementiacouncil.org |

b) Non-condition-specific websites

| Number | Organisation | website |
| --- | --- | --- |
| 56 | AARP (USA) | aarp.org |
| 57 | Age UK | ageuk.org.uk |
| 58 | Ageing well Without Children (UK) | awwoc.org |
| 59 | Association of British Neurologists | theabn.org |
| 60 | British Psychological Society | bps.org.uk |
| 61 | British Society of Gerontology | britishgerontology.org |
| 62 | Centers for Disease Control and Prevention (USA) | cdc.gov |
| 63 | Centre for Ageing Better (UK) | ageing-better.org.uk |
| 64 | Health Policy Partnership (UK) | healthpolicypartnership.com |
| 65 | International Federation on Ageing | ifa.ngo |
| 66 | Joseph Rowntree Foundation (UK) | jrf.org.uk |
| 67 | Meaningful Ageing Australia | meaningfulageing.org.au |
| 68 | Neurological Alliance (UK) | neural.org.uk |
| 69 | NHS England | england.nhs.uk |
| 70 | Royal College of General Practitioners (UK) | rcgp.org.uk |
| 71 | Royal College of Occupational Therapists (UK) | rcot.co.uk |
| 72 | Royal College of Psychiatrists (UK) | rcpsych.ac.uk |
| 73 | The Health Foundation (UK) | health.org.uk |
| 74 | The King's Fund (UK) | kingsfund.org.uk |
| 75 | UK Government | service.gov.uk |
| 76 | World Health Organization | who.int |
